# Shared Antigen-specific CD8^+^ T cell Responses Against the SARS-COV-2 Spike Protein in HLA-A*02:01 COVID-19 Participants

**DOI:** 10.1101/2020.05.04.20085779

**Authors:** William Chour, Alexander M. Xu, Alphonsus H.C. Ng, Jongchan Choi, Jingyi Xie, Dan Yuan, Diana C. DeLucia, Rick A. Edmark, Lesley C. Jones, Thomas M. Schmitt, Mary E. Chaffee, Venkata R. Duvvuri, Kim M. Murray, Songming Peng, Julie Wallick, Heather A. Algren, William R. Berrington, D. Shane O’Mahony, John K. Lee, Philip D. Greenberg, Jason D. Goldman, James R. Heath

## Abstract

We report here on antigens from the SARS-CoV-2 virus spike protein, that when presented by Class I MHC, can lead to cytotoxic CD8^+^ T cell anti-viral responses in COVID-19 patients. We present a method in which the SARS-CoV-2 spike protein is converted into a library of peptide antigen-Major Histocompatibility Complexes (pMHCs) as single chain trimers that contain the peptide antigen, the MHC HLA allele subunit, and the β-2 microglobulin subunit. This library is used to detect the evolution of virus-specific T cell populations in four COVID-19 study participants two of which share one HLA allele, and the other two a second HLA allele, at two time points over the initial course of infection. HLA-matched participants exhibit similar virus-specific T cell populations, but very different time-trajectories of those populations. This strategy can be used to track those virus-specific T cell populations over the course of an infection, thus providing deep insight into the SARS-CoV-2 immune system trajectories observed in different COVID-19 patients.

## Introduction

In December 2019, an outbreak of unexplained pneumonia was recognized to be associated with an open market in Wuhan, China and reported to the World Health Organization (WHO) by mid-January. The causative agent of the disease outbreak, termed COVID-19, was noted to be a novel coronavirus.^1^ Clinical features include fever, cough and dyspnea with a proportion of persons progressing to viral pneumonia and acute respiratory distress syndrome.^2,3^ A high mortality rate was noted in persons requiring mechanical ventilation.^4,5^ A larger case-series of >70,000 participants from the CDC China reported that the majority of cases (81%) were mild, with 14% requiring hospitalization, 5% requiring intensive care and an associated 2.3% proportional mortality.^6^ The case fatality rate is estimated at between 1% and 5%, but varies with age and comorbidities. Elderly individuals are particularly at risk, with the China experience demonstrating 8% mortality in persons aged 70 – 79, and 15% in those aged 80 or older.^6^

Little is known about specifics of host T cell defense responses against SARS-CoV-2, although it has been reported that the severity of the COVID-19 infection correlates with a depletion in both CD4^+^ and CD8^+^ T cell populations, as well as an upregulation of exhaustion markers in those remaining T cells.^7^ One important step towards tracking down the root causes of such immune dysfunction is to identify those T cell populations that are specifically activated by exposure to SARS-CoV-2 antigens. Here we describe a set of measurements in which we identify CD8^+^ T cell antigens arising from the SARS-CoV-2 spike protein as presented by the most common HLA Class I allele A*02:01. Antigen-specific T cell populations were identified from peripheral blood mononuclear cells (PBMCs) isolated from serial draws from two COVID-19 study participants. Similar antigen-specific T cell populations, but very different time-dependent population evolutions, were observed in the two participants.

## Results

Here we describe the prediction of putative SARS-CoV-2 T cell antigens, the validation of experimental reagents, and the measurement process flow.

Using the NetMHC4.0 binding prediction algorithm,^8,9^ we tested all possible 9-mer to 11-mer spike protein epitopes, and identified 96 peptides that bind to the HLA-A*02:01 allele with 500 nM or stronger binding affinity (Supplementary Table S1). These matched reasonably well with published lists of putative antigens.^10–12^ We then utilized standard plasmid cloning techniques, followed by mammalian cell transfection and expression, to build a library of soluble, secreted peptide-major histocompatibility complexes (pMHCs), in which each library element is a single chain trimer (SCT) that contains the peptide antigen covalently linked to the β-2 microglobulin subunit linked to the HLA subunit (Fig. 1A).^13–16^ These SCT libraries can be biotinylated and incorporated into standard pMHC tetramer scaffolds. The SCT tetramers can then be assembled onto the surface of magnetic nanoparticles (to form pNP libraries) for hemocytometry fluorescence microscopy assays (Fig. 1B), similar to our previous reports,^17,18^ or they can be used for standard flow cytometry and fluorescence-activated cell-sorting (FACS) assays (Fig. 1C). For the analysis of COVID-19 participant samples, the pNP libraries have the advantage that all analysis is done in solution, thus avoiding potential risks from aerosolized COVID-19 participant biospecimens.

**Fig. 1.**
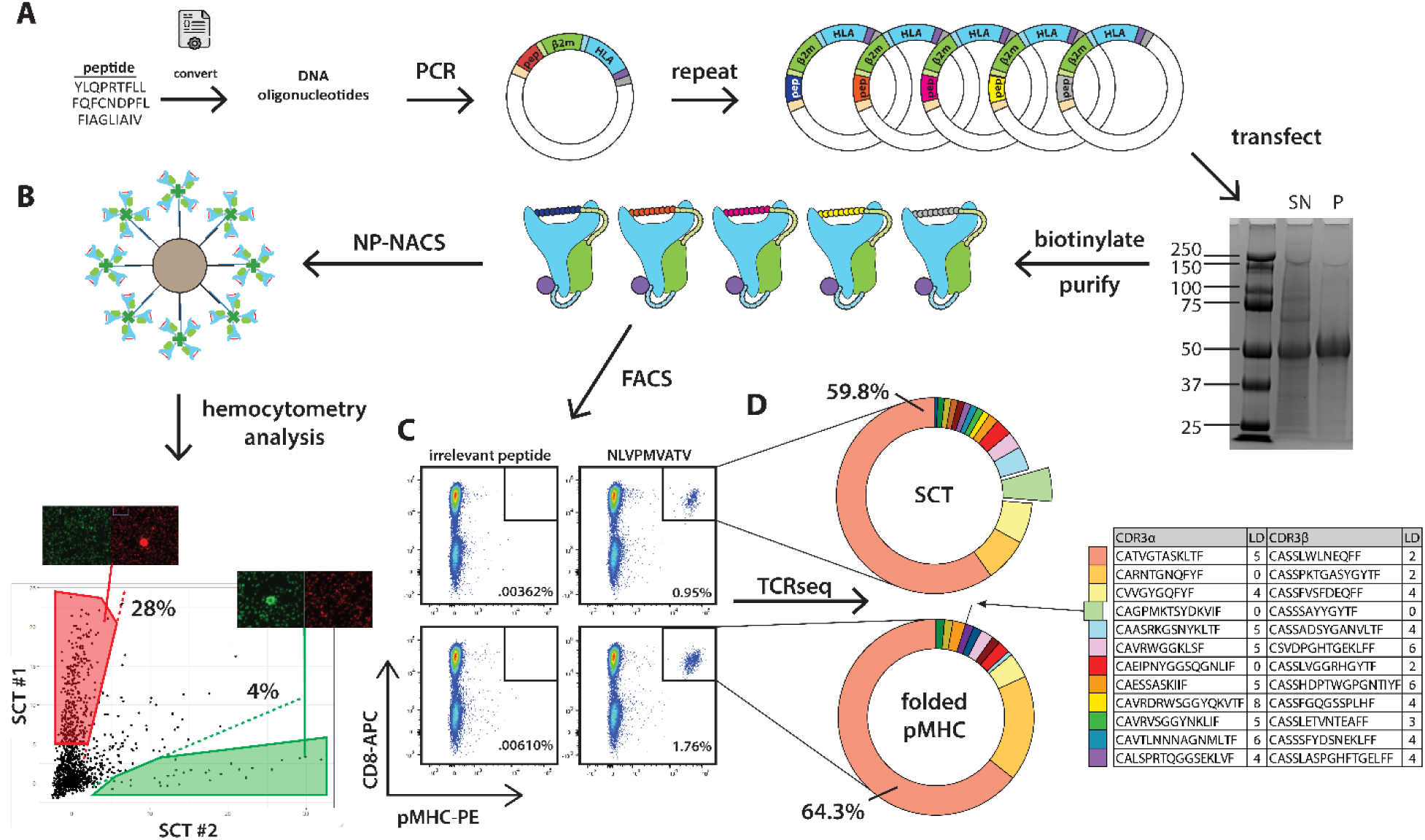
The construction and demonstration of single-chain trimer libraries for antigen-specific T cell capture and analysis. **A**. Process flow for SCT library development. A list of peptide candidates are converted into DNA oligonucleotides for cloning into a plasmid template. The SCT plasmid library is transfected into HEK293 mammalian cells for 4-day incubation and protein expression. Expression of WT1 (RMFPNAPYL) SCT is shown in SDS-PAGE reducing conditions (right). SN, supernatant from transfection; P, HisTag-purified SCT. Numbers on gel denote kDA mass of protein ladder bands. Expressed SCTs are biotinylated for assembly into pMHC tetramers which are then either assembled onto fluorophore-labeled nanoparticles to make a pNP library (B) or used as fluorophore-labeled tetramers for flow cytometry type assays (C). **B**. Use of an SCT-based pNP library to capture antigen-specific T cell populations, with measurements done on a hemocytometer chip using fluorescence microscopy. The data shown are from a 4-color analysis of PBMCs containing MART-1 specific CD8^+^ T cells spiked in as a positive control. A second color is used for a live cell stain, while the 3^rd^ and 4^th^ colors are for two SCT library elements. The cells, when surrounded by a single color pNP, appear as fluorescent donut-shaped structures in the micrograph. **C**. Comparison of a CMV antigen-presenting SCT library element and the analogous pMHC structure prepared via refolding of the separate components. The flow cytometry assays show a similar labeling efficiency for both the SCT and the folded pMHC. **D**. TCR CDR3α/β gene sequencing analysis of CMV-specific T cells captured by SCT and folded pMHC. Pie charts reveal similar clonotypes captured using both reagents. The light green clonotype (offset wedge in both charts and indicated by arrow for folded pMHC) is seen at higher frequency using the SCT tetramer, and corresponds to published CMV-specific CDR3α and CDR3β chains. Table (right) contains CDR3α and CDR3β sequences of the twelve most frequently captured clonotypes from SCT tetramer along with their Levenshtein distance to publicly reported CMV-specific clonotypes from VDJdb.^23^

Our approach to pMHC SCT library synthesis utilized mammalian protein expression systems, as previously reported for individual pMHC constructs.^19^ The application of the platform for library assembly of a diverse set of antigens greatly extends the utility of these reagents. In order to validate our library approach, we first expressed a small SCT library consisting of bacterial and viral antigens (Supplementary Table S2) bound to the MHC by a cysteine-modified glycine linker (Fig. S1A).^20–22^ The transfection experiment to produce these SCTs was performed in triplicate, to demonstrate that SCT protein yield was highly variable in a consistent, antigen-dependent manner (Fig. S1B). Unique to the SCT platform is the expression of pMHCs containing N-linked glycosylated peptides, as evidenced by the increased mass of the library element KLIANNTRV (Fig. S1A). To validate the functionality of SCTs, we expressed an SCT that incorporates an immunogenic peptide derived from human cytomegalovirus hCMV-pp65 protein (NLVPMVATV) for use in antigen-specific T cell capture and analysis (Fig. S1). For A*02:01 haplotype participants with pre-exposure to CMV, T cell populations with specificity to this antigen are readily detected. We compared antigen-specific T cell capture using this CMV SCT construct and its folded pMHC counterpart as a positive control. Flow cytometry results (Fig. 1C) indicated similar efficiencies of cell labeling from both reagents when applied to a healthy A*02:01 donor sample with known reactivity to the CMV epitope. The FACS-sorted cells were sequenced using 10X T-cell receptor (TCR) sequencing technologies, demonstrating that the spectrum and frequency of antigen-specific clones captured by the two reagents are similar. As seen in the table in Figure 1D, Levenshtein distances (LD) of the CDR3α and CDR3β chains against a public database (VDJdb) were low, indicating high similarity between our detected CMV-specific TCR chains and those previously reported.^23^ Two paired clones (red and light orange wedges in the pie graphs of Fig. 1D) contained CDR3α chains that exactly match those found in the literature (LD = 0).^24,25^ An additional clone (light green wedge) contained an α/β pair for which both chains have been reported as specific to the CMV antigen,^24,26^ and was captured by the SCT pMHC variant at a 10fold higher frequency than by the folded pMHC. These data gave us confidence that the SCT platform could be used to present a library of putative SAR-CoV-2 antigens for antigen-specific T cell capture. We further assessed previously published cysteine linker SCT designs and an intra-pocket mutation (H74L) to identify an optimal SCT variant that would bind to T cells with cognate TCRs (Fig. S2A and S2B).^21,27^ In addition, disulfide bonds designed to stabilize α1 and α2 helices of the HLA peptide binding pocket^28–30^ (Y84C and A139C) were incorporated to generate a new disulfide SCT (DS-SCT) template. Flow cytometry assays were performed in which six SCT mutants containing the A*02:01 Wilms Tumor 1 (WT1) antigen (RMFPNAPYL) were used to target T cells transduced with the cognate C4 TCR (Fig. S2C).^31^ WT1 DS-SCT tetramers displayed the best performance against the WT1-specific C4 TCR, capturing up to 94% of the total cell population (Fig. S2D). This design was implemented as the standard template for spike protein derived SCTs.

Figure 2 illustrates how the spike protein library is composed of predicted A*02:01 antigens that broadly sample the different domains of the protein. In Figure 2A, the top color-coded vector diagram illustrates the different domains of the protein, including the literature-described N-Terminal Domain (NTD) (blue) and Receptor Binding Domain (RBD) (green) and the Heptad Repeat (HR) 1 and 2 regions (yellow and magenta).^32^ The small squares below the vector diagram show how the putative antigens map onto the different regions of the spike protein. In Figures 2B & 2C, we provide SCT epitope expression data for 22 of these putative antigens (see Fig. S3A for full dataset). Note, for example, that #’s 2, 5, and 12–14 are strongly expressed, but that there are variable degrees of expression for most SCT epitopes. The majority of the SCT epitopes with little or no expression (for example #3 and #9) can be traced to similar regions of the protein, such as the hydrophobic transmembrane region near the C-terminus (Fig. 2A). Out of 96 attempted SCTs, 60 yielded usable product, of which we tested the top 30 (ranked by relative SCT epitope expression). A comparison of the relative expression yield for each SCT demonstrated no relationship with predicted binding affinity (Fig. 2C & S3B). Peptide 9-mers and 10-mers were more likely to result in expressed SCT epitopes, but the relationship with yield was non-significant (Fig. S3C). The full list of putative antigens and their relative expression yields as SCTs is provided in Supplementary Table S1.

**Fig. 2.**
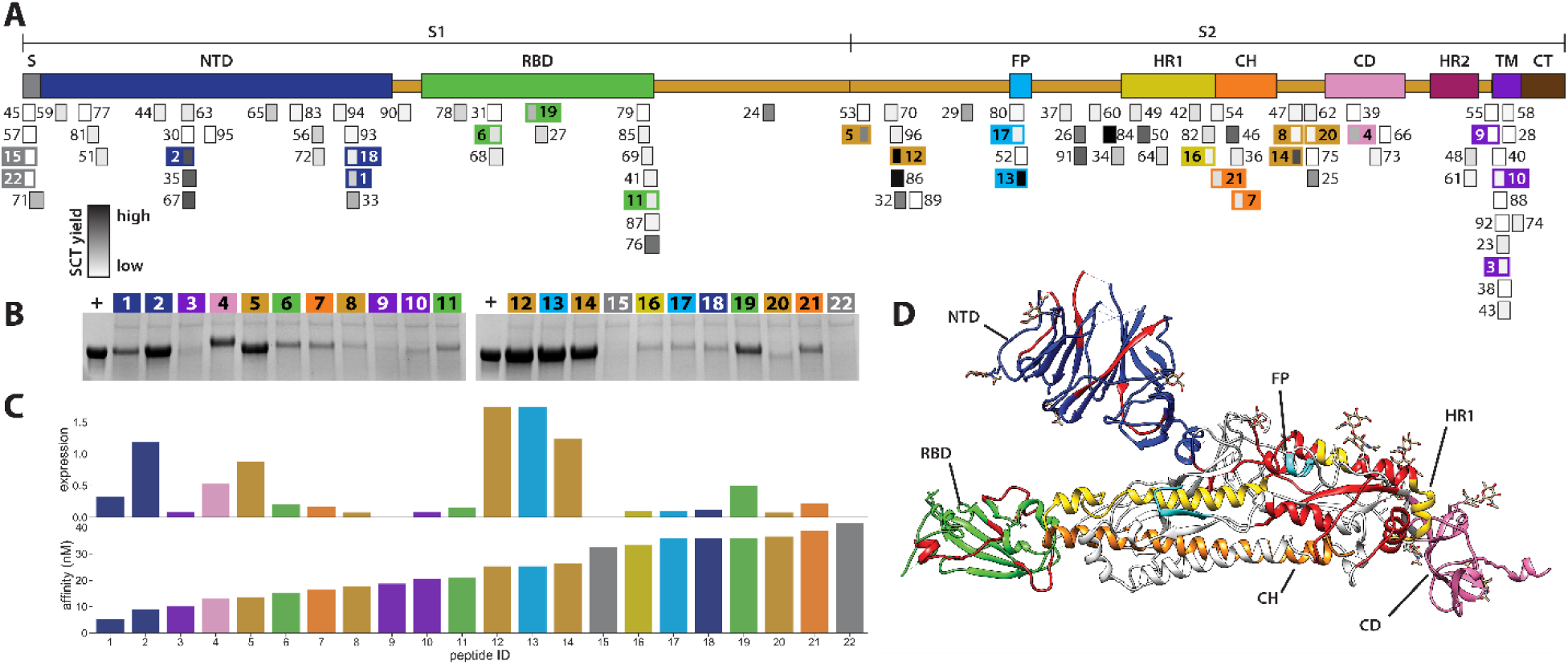
The distribution of putative HLA-A*02:01 T cell antigens throughout the SARS-CoV-2 spike protein. **A**. Schematic of the domains of the SARS-COV-2 spike protein. S, signal sequence; NTD, N-terminal domain; RBD, receptor binding domain; FP, fusion peptide; HR1, heptad repeat 1; CH, central helix; CD, connector domain; HR2, heptad repeat 2; TM, transmembrane domain; CT, cytoplasmic tail; subunits denoted by S1 and S2. Boxes shaded in grayscale denote relative transfection yield of SCT protein for peptide corresponding to the box’s overlapped spike protein region. Numbers correspond to peptide ID (see Supplementary Table S1) and are indexed in descending order of predicted binding affinity. **B**. Expression of a subset of spike epitope SCT proteins. SCT yield for each epitope depicted by SDS-PAGE under reducing conditions. Gel lane number indicates peptide ID, with background color matching domain origin. +, HisTag-purified WT1 SCT, as mass reference and positive control. **C**. Barplots comparing relative SCT expression quantified against positive control lanes and NetMHC4.0 predicted affinity for each peptide from the subset in (B). **D**. Crystal structure of a monomer unit from the spike protein.^32^ Domain colors are matched to those of the schematic in (A); S1 and S2 subunit backbones in white. Amino acids containing the 30 tested epitopes tested in Fig. 2 are colored in red.

### Case Summaries

Participant #2 (InCov002) is a male in his 30s who was previously healthy except for childhood asthma. He presented 5 days after onset of fever, cough and progressive dyspnea. He had myalgias, occasional emesis, and hyperesthesia of the scalp. On arrival, nasal swab for SARS-CoV-2 PCR was positive, and chest X-ray showed patchy bilateral opacities (Fig. S4A). He was initially treated with hydroxychloroquine and azithromycin, but developed progressive acute hypoxemic respiratory failure. On hospital day two (HD2) he required increasing supplemental oxygen and was placed on a high-flow nasal cannula (HFNC) at 30L/min. Based in part on worsening hypoxia and doubling of C-reactive protein (CRP) (Fig. S4C), on HD3 he was diagnosed with cytokine release syndrome (CRS) and treated with tocilizumab, an anti-IL-6 receptor monoclonal antibody^33^ and remdesivir (nucleotide analog inhibitor of SARS-CoV-2 RNA-dependent RNA polymerase).^34^ Cytokine testing prior to tocilizumab showed elevated IL-6. On HD4 he required mechanical ventilation and needed norepinephrine to support mean arterial pressure > 65 mmHg. He received subsequent doses of tocilizumab on HD5 and HD6 receiving a total dose of 24 mg/kg in 3 doses. Remdesivir was stopped on HD7 for elevation of liver transaminases (Fig. S4C). He was extubated on HD9 and discharged home on HD12 to telehealth monitoring.

Participant #5 (InCov005) is a male in his 70s who has hypertension treated with angiotensin receptor blockers (ARBs), and a history of asbestos exposure and lung nodules. He has moderate chronic obstructive pulmonary disease (COPD) and chronic bronchitis, with a 40 pack-year smoking history, quit 10 years ago, and recent postbronchodilator FEV_1_ 2.05L (65% predicted) (Fig. S5C). He was treated with multimodal therapy for COPD, including inhaled and intermittent systemic steroids (Fig. S5D). Three weeks prior to arrival, he saw his pulmonologist and started a steroid taper, which finished 8 days prior to presentation. He developed fever and worsening cough 1 day prior to presentation in the emergency room, with other symptoms that included insomnia and anorexia. On arrival, nasal swab for SARS-CoV-2 PCR was positive, and CT angiogram of the chest showed patchy ground-glass opacities peripherally with superimposed moderate to severe emphysematous changes (Fig. S5A). By HD2, he required supplemental O_2_ by HFNC 30 L/min to maintain SpO_2_ > 92% and was started on remdesivir for a 10-day course. On HD3, due to increasing hypoxia and CRP increased to 306 mg/L, he was diagnosed with CRS and treated with tocilizumab. He continued to worsen with higher oxygen requirements by HD5, and received 2 further doses of tocilizumab, for total dose of 24 mg/kg in 3 doses. Cytokine testing after tocilizumab completion showed persistent elevation in IL-6 (Fig. S5B). He avoided intubation, but required HFNC though HD17, and was discharged to home on HD19, newly requiring home supplemental O_2_.

We analyzed for populations of SARS-CoV-2 antigen-specific T cells from a single healthy A*02:01 haplotype control, and two COVID-19 study participants (InCov002 and InCov005) at the time of initial clinical diagnosis (T1), and at a time point 5 days later for InCov002 and 4 days later for InCov005 (T2). Metadata on these participants is provided in Supplementary Table S3, and full HLA haplotyping of these participants is provided in Supplementary Table S4. Blood was collected and analyzed following participant consent using IRB-approved methods and procedures.

Detected spike protein antigen specific CD8^+^ T cell population frequencies for participants InCov002 and InCov005 are shown in Figure 3. Note the differing y-axis scales for these data. For participant InCov005 at the T1 time point, CD8^+^ T cell analysis revealed 14, 66, 13, 33, 6, and 13 cells per 10^4^ cells were specific to six antigens derived from NTD, RBD, FP, CH, and S2 domains (see Fig. 3 x-axis for sequences and domain-mapped locations). CD8^+^ T cell populations specific to these antigens were still observed at T2, but with an average 3-fold drop in frequency. For participant InCov002 at the T1 time point, CD8^+^ T cell analysis shows that five of the six populations seen in participant InCov005 were detected above 1–2 counts per 10^4^ cells, although at slightly lower frequencies relative to participant InCov005. Vertical dashed lines on the graph show these shared responses between the participants. The kinetic trend for participant InCov002 is opposite that of participant InCov005. At the later time point, the five populations exhibited an approximately 2-fold increase. Notably, T cells specific to the RBD, FP, and CH/S2 derived antigens are detected in the healthy donor, but at 10–20-fold lower frequencies than in the COVID-19 participants. This is consistent with published reports that similar haplotype healthy donors can exhibit measurable, but low frequencies of antigen-specific CD8^+^ T cell populations relative to what is seen in diseased individuals.^35^ It may also imply a level of genetic encoding of the T cell responses shared among similar haplotype individuals. Additional HLA-matched control participants will need to be investigated to resolve this.

**Fig. 3.**
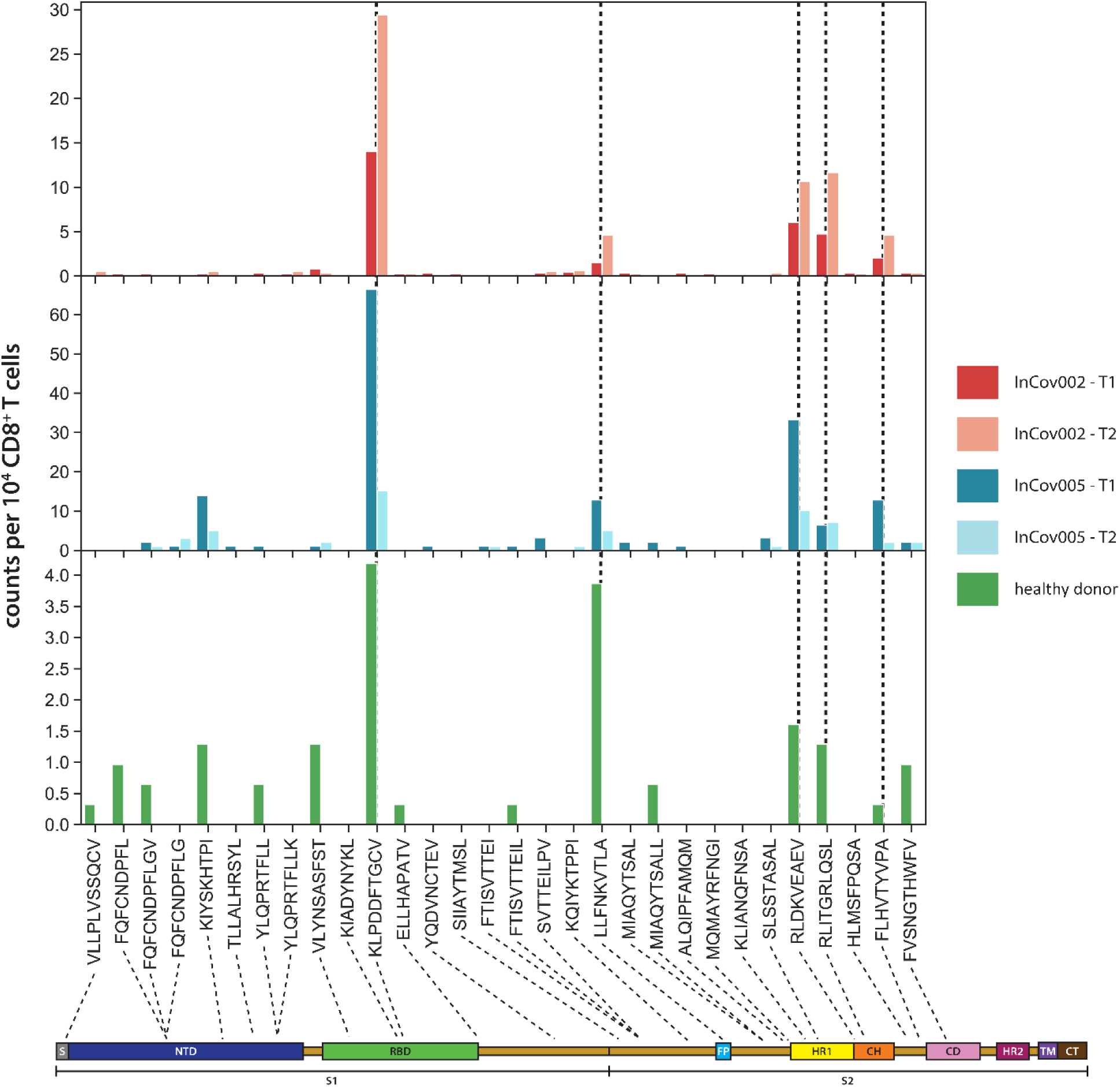
T cell populations detected in serial draws from two HLA-A*02:01 COVID-19 participants and healthy HLA-A*02:01 control. The antigens are plotted according to their position on the spike protein, from the N-to-C termini. The dashed vertical lines within the plot are visual aids to highlight those populations seen in both COVID-19 participants at both time points. Dashed lines under each peptide denote its position along the domain map of spike protein. Note the different y-axis scales on the three plots.

## Discussion

The strong correlation between spike protein antigen-specific T cell populations in both participants InCov002 and InCov005 indicates that the overlapping five antigens may be immunodominant, and that such T cell populations may also be predicted to be seen in other COVID-19 patients with the same HLA-A*02:01 haplotype. The time-dependent behaviors of these T cell populations may reflect differing anti-SARS-CoV-2 immune trajectories in these two participants, leading to different eventual clinical outcomes.

The spike protein derived SCT library tested here was limited. First, only 60 (out of 96 putative antigens) could be expressed as SCTs. From these 60, only the 30 most highly expressed SCTs were tested. While these SCTs broadly sampled the range of predicted antigen-MHC affinities, the majority of T cells detected were specific to antigens characterized by a binding affinity stronger than 100 nM (Fig. S6). Thus, it is likely that a thorough search would reveal additional immunogenic antigens. A second bias is in the A*02:01 allele, which does not permit sampling of certain parts of the spike protein, such as the S1/S2 cleavage domain. That domain does yield putative antigens that are sampled by other HLA alleles, and next steps will be to create additional SCT libraries for different HLA alleles. One possibility is that the different immunogenic epitopes of the spike protein confer differing levels of immune protection, based upon factors such as epitope-dependent genetic stability. Understanding the scope of immunodominant epitopes across common and rare HLA haplotypes will be important to understand population response to peptide vaccines.

A notable difference is the increasing CD8^+^ T cell response in participant InCov002 versus the declining CD8^+^ T cell response in participant InCov005, as indicated by the CD8^+^ T cell frequency counts for immunodominant responses increasing and decreasing, respectively. While both participants received remdesivir and aggressive dosing of tocilizumab (24 mg/kg each), and both developed acute hypoxemic respiratory failure, participant InCov005 never required mechanical ventilation, despite advanced age and a much worse baseline pulmonary status due to COPD. In this case, perhaps the prior steroids and age-related immunosenescence were helpful in tamping the more vigorous immune response seen in participant InCov002 that led to CRS, hypotension and need for mechanical ventilation.

## Methods

### Study design

The objective of this study was to explore the role of the antigen-specific adaptive immune response against SARS-nCOV-2. To this end, pMHCs were designed in the form of a single chain trimer (SCT) comprising a candidate SARS-nCoV-2-derived spike protein epitope, β-2 microglobulin subunit of the MHC, and the human leukocyte antigen (HLA) subunit of the MHC. The optimized platform was utilized to express approximately 60 viable SCT constructs against the spike protein. We then incorporated 30 of these SCT constructs as tetramers into our nanoparticle nucleic acid cell sorting (NP-NACS) system to generate high-avidity TCR capture agents. Last, NP-NACS was applied toward the identification and analysis of antigen-specific T cells derived from blood draws of two COVID-19 participants, as well as from a healthy donor PBMC sample.

### Study approval

All human samples (blood) were obtained after approval from the Providence St. Joseph’s Health IRB) and participant-written informed consent, in accordance with 45 CFR 46.

### Peptide selection

The FASTA protein sequence of the spike surface glycoprotein from severe acute respiratory syndrome coronavirus 2 isolate Wuhan-Hu-1 (NCBI Reference Sequence: YP_009724390.1) was submitted to the NetMHC4.0 server (http://www.cbs.dtu.dk/services/NetMHC/) to predict binding affinities of all possible 9-mer, 10-mer, and 11-mer epitopes with HLA-A*02:01 allele. The output of predicted binding affinities was indexed according to bind strength, and the top 96 peptides (approximately 5 to 500 nM affinity) were selected for development into SCT plasmid constructs.

### SCT plasmid design and production

Single-chain trimer constructs were produced according to design parameters previously reported with conventional plasmid cloning techniques.^13,36^ The pcDNA mammalian expression vector (Thermo Fisher Scientific) encoding (MART-1)-β2m-(HLA-A*02:01) was used as starting material for the assembly of additional SCT constructs. It contains the MART-1 self-antigen (ELAGIGILTV) to serve as an oligonucleotide placeholder. Oligonucleotides containing desired insert antigens and other modifications reported in this article were substituted into the plasmid via standard cloning procedures as reported by others.^37,38^ The constructs were transfected into HEK293-derived cell lines with polyethylenimine (Thermo Fisher Scientific). The solubilized SCTs were biotinylated by BirA enzyme (Avidity), and subsequently extracted from transfection medium by purification with an immobilized nickel column. SCTs were resuspended into PBS with 20% glycerol for long-term storage and application.

### SCT protein expression and characterization

A 15 µl solution containing 3:1 mix of transfection supernatant and Laemmli buffer with 10% β-mercaptoethanol was denatured at 100°C for 10 minutes, and subsequently loaded into Bio-Rad Stain-Free gels for SDS-PAGE (200V, 30 minutes). A purified WT1 SCT sample in 20% glycerol PBS solution (2 µg) was run in each gel to serve as a positive control and reference for relative protein yield calculation. Images were obtained using a Bio-Rad ChemiDoc MP gel imaging system (manual settings: 45 seconds UV activation, 0.5 second exposure). Band detection and quantification with Python scripts were utilized to compare relative SCT yields. The accuracy of this approach was measured by SDS-PAGE of titrated, pre-quantified samples of purified SCTs, to demonstrate a 99% correlation between true protein A280 concentration (as measured by NanoDrop 8000 Spectrophotometer) and quantified relative band intensity (data not shown).

### Production of cysteine-modified streptavidin-DNA (SAC-DNA) conjugates

The SAC-DNA conjugate was produced following a previously published protocol.^39^ Briefly, SAC was first expressed from the pTSA-C plasmid containing the SAC gene (Addgene).^40^ Before conjugation to DNA, SAC (1 mg/ml) was buffer exchanged to PBS containing Tris(2-Carboxyethyl) phosphine hydrochloride (TCEP, 5 mM) using Zeba desalting columns (Pierce). Then 3-N-Maleimido-6-hydraziniumpyridine hydrochloride (MHPH, 100 mM, Solulink) in DMF was added to SAC at a molar excess of 300:1. In the meantime, succinimidyl 4-formylbenzoate (SFB, 100mM, Solulink) in DMF was added to 5’-amine modified ssDNA (500 µM) in a 40:1 molar ratio. After reacting at rt for 4 hours, MHPH-labeled SAC and SFB-labeled DNA were buffer exchanged to citrate buffer (50 mM sodium citrate, 150 mM NaCl, pH 6.0), and then mixed at a 20:1 ratio of DNA to SAC to react at rt overnight. SAC-DNA conjugate was purified using the Superdex 200 gel filtration column (GE health) and concentrated with 10K MWCO ultra-centrifuge filters (Millipore).

### COVID SCT pNP library construction

Streptavidin-coated NPs (500 nm radius, Invitrogen Dynabeads MyOne T1) were prepared according to the manufacturer’s recommended protocol for biotinylated nucleic acid attachment. These NPs were mixed with barcoded biotin-ssDNA (100 µM) at 1:20 volume ratio to obtain NP-DNA. Excess DNA was removed by washing the NPs three times. In parallel, the SCT monomer library was added to SAC-DNA at a 4:1 ratio to form the SCT tetramer-DNA. To generate fluorescent pNPs, equimolar amounts (in terms of DNA ratio) of NP-DNA and pMHC tetramer-DNA were hybridized at 37°C for 20 min, along with 0.25 µl of 100 µM ssDNA bound to AlexaFluor 750, AlexaFluor 488, or Cy5 (IDT-DNA), and washed once with buffer (0.1% BSA, 2 mM MgCl_2_ PBS). The use of three dyes allows for multiplexing of up to three unique antigen pNPs per analysis. Typically, each NP-barcoded NACS analysis of <100,000 cells uses 2.5 µL of stock NPs (28.2 million particles total) per library element.

### Preparation and isolation of CD8^+^ T cells from PBMC suspensions

Whole blood specimens were collected from participants infected with the novel coronavirus SARS-CoV-2 at two different time points, T1 time point was after providing informed consent, HD3 for InCov002 and HD2 for InCov005, and T2 time point was 5 days later for InCov002 and 4 days later for InCov005. Peripheral blood mononuclear cells (PBMCs) were isolated and cryopreserved by Bloodworks Northwest (North Seattle Donor Center, Seattle). PBMCs were thawed and incubated in RPMI 1640 media supplemented with 10% FBS and IL2 (100 U/mL) for overnight recovery at 37°C, 5% CO_2_. Recovered cell viability was measured at >95% for all samples.

CD8^+^ T cell population was negatively selected using the CD8^+^ T Cell Isolation Kit (Miltenyi biotec, Bergisch Gladbach, Germany). Briefly, recovered cells were incubated with a biotinylated antibody cocktail that captures non-CD8^+^ cells in PBMCs followed by streptavidin-coated microbeads. The untouched CD8^+^ T-cells were separated in a 15 mL Falcon tube using a LS column. The centrifuge tube containing CD8^+^ T-cells was then centrifuged at 500 g for 5 minutes and the pellet was resuspended in PBS buffer. The number of sorted CD8^+^ T cells were counted by a hemocytometer.

For the multiplex cell labeling, CD8^+^ T-cells were individually stained with Calcein Blue, AM (Thermo Fisher Scientific) or CellTracker™ Orange CMRA Dye (Thermo Fisher Scientific) at the concentration of 4 µM and 400 nM, respectively. After incubation for 10 minutes at 37°C under 5% CO_2_, cells were washed twice with PBS and resuspended in a cell suspension buffer (0.1% BSA, 2 mM MgCl_2_ in PBS).

### Identification of antigen-specific CD8^+^ T cells by NP-NACS

The pNP library was combined into groups of 3 pNPs, with each pNP element in the group stained with one of three barcode dyes. From each pNP group, 7.5 µl was incubated with each aliquot of stained CD8^+^ T cells at rt for 30 minutes. Antigen-specific cells were enriched by magnet pulldown and re-suspended into 6 µl of 0.1% BSA 2 mM MgCl_2_ PBS buffer. Captured cells were then loaded into a 4-chip disposable hemocytometer (Bulldog-Bio). The entire area in the hemocytometer chip was imaged to obtain the total pulldown cell number. Identification of antigen-specific T cells, including the detection and exclusion of non-specific binding events, was conducted with scripts written in R programming language.

## Data Availability

Please contact the ISB data manager for release of data.

## Acknowledgments

We acknowledge the Parker Institute for Cancer Immunotherapy, the Andy Hill CARE Fund, the Wilke Family Foundation, the M.J. Murdock Charitable Trust, the Swedish Foundation, Merck, and the Biomedical Advanced Research and Development Authority (BARDA) for their generous support of this work.

